# Cause of Death by Race and Ethnicity in Minnesota Before and During the COVID-19 Pandemic, 2019-2020

**DOI:** 10.1101/2023.03.09.23287048

**Authors:** Madelyn J. Blake, Nicholas A. Marka, Clifford J. Steer, Jonathan I. Ravdin

**Author notes:** **Corresponding Author:** Madelyn J. Blake, University of Minnesota, Minneapolis, MN, 55347. Footnote: This manuscript is dedicated to the memory of Jonathan I. Ravdin, scholar, mentor, physician-scientist and friend. **Author Contributions:** Madelyn Blake had full access to all the data in the study and takes responsibility for the integrity of the data and the accuracy of the data analysis. Concept and Design:* Blake, Ravdin, Marka. Acquisition, Analysis, and Interpretation of Data:* Blake, Ravdin, Marka. Drafting of the Manuscript:* Blake, Marka, Steer. Review and Revisions of the Manuscript:* Blake, Marka, Steer. Statistical Analysis:* Blake, Marka. Obtained Funding:* Ravdin. Supervision:* Ravdin, Steer.

## Abstract

**Objectives:** To measure changes in cause of death dynamics in 2019 and 2020 and the relationship between concurrent occurrence of the COVID-19 pandemic and mortality outcome by race and ethnicity.

**Patients and Methods:** We used resident mortality data from the Minnesota Department of Health (MDH) to conduct retrospective statistical analysis of deaths in Minnesota in 2019 relative to 2020 to assess changes in mortality in a pre-pandemic and pandemic period.

**Results:** COVID-19 strongly contributed to ethnicity-related mortality disparities in Minnesota. Not only was there a greater proportion of COVID-19 decedents within the Black and Hispanic populations, but their average decedent age was markedly lower relative to the White population. The Black population experienced a disproportionate increase in decedents with a 34% increase during 2020 compared to 2019.

**Conclusions:** This retrospective analysis of death dynamics and mortality outcomes in Minnesota from 2019 to 2020 demonstrated an increase in adverse mortality outcomes relative to the pre-pandemic period that disproportionately impacted Black and Hispanic minority populations. Access to non-pharmaceutical interventions combating COVID-19 infection in Black and Hispanic communities should be expanded in Minnesota.

## INTRODUCTION

The adverse effects of coronavirus disease of 2019 (COVID-19) on racial and ethnic inequalities in the United States are well-established in the literature. ^1^ However, the magnitude to which COVID-19 impacted other leading causes of mortality and respective racial or ethnic disparities remains unclear.

Minnesota’s population is overwhelmingly White with 61.1% of its residents identifying as White, 12.4% identifying as Black, and 18.7% identifying as Hispanic or Latino of any race.^2^ Despite the lack of diversity in the state overall, the Twin Cities Metro area is becoming increasingly diverse, with 19.7% of its residents reporting a race or ethnicity other than White in 2018. Moreover, there are similar labor force participation rates among Minnesota’s White, Black, and Hispanic residents in the Twin Cities Metro - 72.2%, 72.0%, and 77.6%, respectively. Many industrial sectors within Minnesota’s urban center reflect this diversity, apart from a few essential industries that rely heavily on non-White workers, namely Health Care and Social Assistance, Manufacturing, and Accommodation and Food Services.^4^ The reliance of these essential industries on non-White workers intensified their risk of COVID-19 infection and fostered a greater likelihood of racial and ethnic mortality disparities in Minnesota secondary to the COVID-19 pandemic.

In addition to its economic disparities, Minnesota also possesses substantial racial and ethnic health disparities that predisposed non-White residents to adverse outcomes during the COVID-19 pandemic. Black and Hispanic residents possess depressed health outcomes relative to White residents in Minnesota, particularly regarding cardiovascular and diabetes care.^5^ These adverse outcomes are due to the disproportionate poverty and lack of health insurance among non-White residents in Minnesota. The proportion of Black and Hispanic residents below the poverty level in 2019 was 20.7% and 11.2% respectively, as opposed to only 7.0% for Minnesota’s White residents.^6^ Moreover, Minnesota’s Black and Hispanic residents are far less likely to have access to healthcare. As example, in 2020, individuals that identified as Black alone possessed a community uninsured rate of 7.5% and for individuals identifying as Hispanic or Latino of any race, the community uninsured rate was 17.1%. Minnesota’s residents identifying as White alone possessed a community uninsured rate of only 3.6% in comparison.^7^ Collectively, these health inequities also predisposed Black and Hispanic individuals to more severe complications from COVID-19 infection, namely, acute respiratory distress syndrome (ARDS). ARDS is a life-threatening form of respiratory failure and is a strong indicator of poor prognosis in COVID-19 patients.^8-11^

This study was designed to determine the impact of COVID-19 on cause of death in Minnesota during the onset of the COVID-19 pandemic and investigate the presence of augmented racial and/or ethnic mortality disparities. Specifically, changes in mortality in Minnesota were examined between 2019 and 2020, capturing the year prior to the pandemic and the first year of its onset, and stratified these changes by age and gender in addition to race and ethnicity.

## METHODS

The Minnesota State Institutional Review Board waived requirements for informed consent because no living human subjects participated in the study. Minnesota mortality data were obtained from Minnesota Death Records through a partnership with the Minnesota Department of Health. The obtained data included information on all resident decedents in Minnesota from 2015 to 2020 and included demographic information for each decedent, such as gender, date of birth, and ethnicity alongside pertinent health information. Using the NCHS 113 Selected Causes of Death list, the listed ICD-10 codes for primary cause of death were mapped for each decedent to be consistent with reported CDC mortality statistics. While the data included information on resident decedents from 2015-2020, this study’s analysis focused on deaths in 2019 and 2020. State-level mortality calculations involved all reported Minnesota-resident deaths, including those that were not presently in-state at the time of demise. Mortality data was obtained from the Minnesota Department of Health in May 2021. Population estimates for Minnesota in 2019 and 2020 were obtained from US Census Bureau’s American Community Survey (ACS).^2^

Leading causes of death analysis was conducted by synthesizing decedent population subsets, such as decedent sex or race, and then ranking primary cause of death alongside relative frequency for each respective subset. Mortality analysis involving age utilized the reported decedent age at the time of death, as did mortality analysis involving decedent sex. Cause of death analysis involving race and ethnicity was based on four fixed-response options utilized by the Minnesota Department of Health: Hispanic and non-Hispanic, Black and White populations. For the race and ethnicity mortality analysis, three subsets were created: Hispanic decedents, non-Hispanic White decedents, and non-Hispanic Black decedents. Of note, this paper uses the term “Hispanic” to refer to individuals that self-identify as Latino/Latina, and to maintain consistency with current guidelines for reporting of race and ethnicity.^12^

Mortality analysis involving comorbidities for acute respiratory distress syndrome (ARDS) was conducted by identifying individuals with ARDS comorbidities listed as non-primary causes of death. ARDS is the most severe form of respiratory distress and is a strong indicator of poor clinical prognosis following COVID-19 infection. ARDS comorbidities were broadly defined as: diabetes mellitus, hypertension, and chronic obstructive pulmonary disease, asthma, sepsis, and active malignancy as well as a reported age older than 65.^8-11^

For location-based mortality analysis, the reported decedent location of death was used. Decedent location of death was specified to cities with Minnesota, but this analysis examined decedent death location at the county-level. To analyze the proportion of decedents that expired in the Twin Cities Metro area, a subset was created consisting of the seven counties that compose the Twin Cities Metro: Anoka, Carver, Dakota, Hennepin, Ramsey, Scott, and Washington county.^13^ Analysis pertaining to nursing-home decedents was based on the fixed options provided by the Minnesota Department of Health and described as nursing home resident or non-nursing home resident.

The primary outcome for this study was an assessment of the change in mortality rate in Minnesota between 2019, the period prior to the emergence of the COVID-19 pandemic, and 2020. Mortality alterations in 2020 described the change in cause of death dynamics concurrent with the onset of the COVID-19 pandemic; they do not describe permanent changes to Minnesota mortality dynamics in subsequent years.

Decedent demographics and mortality outcomes were summarized as means for continuous measures, and frequencies and percentages (%) for categorical measures. Comparisons between demographic and clinical groupings between decedents was implemented using a two-sample t-test for continuous outcomes, and Chi-square test for categorical outcomes. All statistical analyses were conducted at the 0.001 significance level using the R statistical software version 4.1.2.^14^

## RESULTS

### Overall Changes in Mortality Between 2019 and 2020

In 2019, Minnesota’s population was 5.6 million – during 2020, the state’s population increased to 5.7 million.^2^ When examining deaths in Minnesota, 45,396 resident decedents were reported in 2019 and 52,192 resident decedents were reported in 2020 and constituted a significant 13% increase from 2019 (95% CI, P < 0.001). In 2020, 5,111 decedents had COVID-19 reported as their primary cause of death, which was approximately 12% of the entire resident decedent population in the state. Overall, COVID-19 ranked as the third leading cause of death in Minnesota for 2020 (Table 1). When examining the change in mortality not directly associated with COVID-19, Minnesota reported 45,396 resident decedents in 2019 and 47,081 in 2020, which composed a significant 4% increase in mortality from 2019 (95% CI, P < 0.001).

**Table 1.** Leading Causes of Death in Minnesota in 2020, Stratified by Age

|  | Age Groups |  |  |  |  |  |  |  |  |  |
| --- | --- | --- | --- | --- | --- | --- | --- | --- | --- | --- |
| Rank | <1 | 5-9 | 10-14 | 15-24 | 25-34 | 35-44 | 45-54 | 55-64 | 65+ | Total |
| 1 | Accidents (Unintentional Injuries): 18 (34%) | Accidents (Unintentional Injuries): 8 (38%) | Accidents (Unintentional Injuries): 12 (33%) | Accidents (Unintentional Injuries): 219 (50%) | Accidents (Unintentional Injuries): 403 (47%) | Accidents (Unintentional Injuries): 331 (31%) | Malignant Neoplasms: 511 (26%) | Malignant Neoplasms: 1658 (36%) | Malignant Neoplasms: 7624 (24%) | Malignant Neoplasms: 10076 (25%) |
| 2 | Congenital Malformations, Deformations, and Chromosomal Abnormalities: 15 (28%) | Malignant Neoplasms: 4 (19%) | Intentional Self-Harm (Suicide): 7 (19%) | Intentional Self-Harm (Suicide): 95 (22%) | Intentional Self-Harm (Suicide): 137 (16%) | Malignant Neoplasms: 185 (17%) | Accidents (Unintentional Injuries): 354 (18%) | Diseases of Heart: 816 (18%) | Diseases of Heart: 7175 (23%) | Diseases of Heart: 8519 (21%) |
| 3 | Assault (Homicide): 5 (9%) | Assault (Homicide): 3 (14%) | Congenital Malformations, Deformations, and Chromosomal Abnormalities: 3 (8%) | Assault (Homicide): 58 (13%) | Malignant Neoplasms: 68 (8%) | Diseases of Heart: 143 (13%) | Diseases of Heart: 319 (16%) | Accidents (Unintentional Injuries): 420 (9%) | COVID-19: 4560 (14%) | COVID-19: 5111 (12%) |
| 4 | Malignant Neoplasms: 5 (9%) | Congenital Malformations, Deformations, and Chromosomal Abnormalities: 2 (10%) | Cerebrovascular Diseases: 2 (6%) | Malignant Neoplasms: 18 (4%) | Assault (Homicide): 63 (7%) | Intentional Self-Harm (Suicide): 128 (12%) | Chronic Liver Disease and Cirrhosis: 169 (9%) | COVID-19: 365 (8%) | Alzheimer Disease: 2552 (8%) | Accidents (Unintentional Injuries): 3334 (8%) |
| 5 | Diseases of Heart: 2 (4%) | *In situ* Neoplasms, Benign Neoplasms, and Neoplasms of Uncertain or Unknown Behavior: 2 (10%) | *In situ* Neoplasms, Benign Neoplasms, and Neoplasms of Uncertain or Unknown Behavior: 2 (6%) | Diseases of Heart: 11 (3%) | Chronic Liver Disease and Cirrhosis: 48 (6%) | Chronic Liver Disease and Cirrhosis: 97 (9%) | COVID-19: 129 (7%) | Chronic Liver Disease and Cirrhosis: 256 (6%) | Cerebrovascular Diseases: 1997 (6%) | Alzheimer Disease: 2580 (6%) |
| 6 | *In Situ* Neoplasms, Benign Neoplasms, and Neoplasms of Uncertain or Unknown Behavior: 2 (4%) | Enterocolitis Due to Clostridium difficile: 1 (5%) | Acute Bronchitis and Bronchiolitis: 1 (3%) | Legal Intervention: 6 (1%) | Diseases of Heart: 48 (6%) | COVID-19: 39 (4%) | Intentional Self-Harm (Suicide): 108 (6%) | Diabetes Mellitus: 230 (5%) | Chronic Lower Respiratory Diseases: 1919 (6%) | Cerebrovascular Diseases: 2277 (6%) |
| 7 | Acute Bronchitis and Bronchiolitis: 1 (2%) | Influenza and Pneumonia: 1 (5%) | Assault (Homicide): 1 (3%) | Chronic Lower Respiratory Diseases: 5 (1%) | Diabetes Mellitus: 22 (3%) | Diabetes Mellitus: 28 (3%) | Diabetes Mellitus: 82 (4%) | Chronic Lower Respiratory Diseases: 226 (5%) | Accidents (Unintentional Injuries): 1546 (5%) | Chronic Lower Respiratory Diseases: 2202 (6%) |
| 8 | Cerebrovascular Diseases: 1 (2%) | N/A | Certain Conditions Originating in the Perinatal Period: 1 (3%) | Congenital Malformations, Deformations, and Chromosomal Abnormalities: 5 (1%) | COVID-19: 14 (2%) | Cerebrovascular Diseases: 23 (2%) | Cerebrovascular Diseases: 73 (4%) | Cerebrovascular Diseases: 170 (4%) | Diabetes Mellitus: 1107 (4%) | Diabetes Mellitus: 1474 (4%) |
| 9 | Certain Conditions Originating in the Perinatal Period: 1 (2%) | N/A | Chronic Lower Respiratory Diseases: 1 (3%) | Diabetes Mellitus: 4 (1%) | Congenital Malformations, and Chromosomal Abnormalities: 9 (1%) | Assault (Homicide): 22 (2%) | Chronic Lower Respiratory Diseases: 38 (2%) | Intentional Self-Harm (Suicide): 118 (3%) | Nephritis, Nephrotic Syndrome, and Nephrosis: 466 (2%) | Chronic Liver Disease and Cirrhosis: 889 (2%) |
| 10 | Complications of Medical and Surgical Care: 1 (2%) | N/A | Complications of Medical and Surgical Care: 1 (3%) | COVID-19: 3 (1%) | Influenza and Pneumonia: 8 (1%) | Congenital Malformations, Deformations, and Chromosomal Abnormalities: 15 (1%) | Influenza and Pneumonia: 23 (1%) | Septicemia: 59 (1%) | Influenza and Pneumonia: 436 (1%) | Intentional Self-Harm (Suicide): 738 (2%) |

The top three leading causes of death in Minnesota during 2019 were malignant neoplasms, cardiovascular disease, and accidents or unintentional injuries (29%, 24%, 8% of the entire decedent population, respectively) (Table 2). In contrast, the three leading causes of death for 2020 were that of malignant neoplasms, cardiovascular disease, and COVID-19 (25%, 21%, and 12% of the entire decedent population, respectively). The observed decrease in malignant neoplasm and cardiovascular disease decedents between 2019 and 2020 was found to be significant (95% CI, P < 0.001). Despite the disproportionate increase in decedents between 2019 and 2020, the average decedent age in Minnesota, 75.8, remained unchanged (95% CI, P < 0.001) (Figure 1).

**Table 2.** Leading Causes of Death in Minnesota for 2019, Stratified by Age

| Age Groups |  |  |  |  |  |  |  |  |  |  |
| --- | --- | --- | --- | --- | --- | --- | --- | --- | --- | --- |
| Rank | <1 | 5-9 | 10-14 | 15-24 | 25-34 | 35-44 | 45-54 | 55-64 | 65+ | Total |
| 1 | Accidents (Unintentional Injuries): 16 (29%) | Congenital Malformations, Deformations, and Chromosomal Abnormalities: 8 (27%) | Accidents (Unintentional Injuries): 14 (35%) | Accidents (Unintentional Injuries): 174 (44%) | Accidents (Unintentional Injuries): 261 (41%) | Accidents (Unintentional Injuries): 244 (27%) | Malignant Neoplasms: 537 (32%) | Malignant Neoplasms: 1686 (41%) | Malignant Neoplasms: 7623 (28%) | Malignant Neoplasms: 10117 (29%) |
| 2 | Congenital Malformations, Deformations, and Chromosomal Abnormalities: 11 (20%) | Accidents (Unintentional Injuries): 6 (20%) | Intentional Self-Harm: 11 (28%) | Intentional Self-Harm (Suicide): 112 (29%) | Intentional Self-Harm (Suicide): 141 (22%) | Malignant Neoplasms: 167 (19%) | Diseases of Heart: 321 (19%) | Diseases of Heart: 745 (18%) | Diseases of Heart: 7099 (26%) | Diseases of Heart: 8347 (24%) |
| 3 | Malignant Neoplasms: 9 (16%) | Malignant Neoplasms: 6 (20%) | Malignant Neoplasms: 5 (13%) | Assault (Homicide): 35 (9%) | Malignant Neoplasms: 60 (9%) | Intentional Self-Harm (Suicide): 134 (15%) | Accidents (Unintentional Injuries): 256 (15%) | Accidents (Unintentional Injuries): 334 (8%) | Alzheimer Disease: 2525 (9%) | Accidents (Unintentional Injuries): 2826 (8%) |
| 4 | Assault (Homicide): 6 (11%) | Assault (Homicide): 3 (10%) | Congenital Malformations, Deformations, and Chromosomal Abnormalities: 4 (10%) | Malignant Neoplasms: 23 (6%) | Assault (Homicide): 46 (7%) | Diseases of Heart: 117 (13%) | Chronic Liver Disease and Cirrhosis: 145 (9%) | Chronic Lower Respiratory Diseases: 225 (5%) | Cerebrovascular Diseases: 2075 (8%) | Alzheimer Disease: 2550 (7%) |
| 5 | Cardiovascular Disease: 4 (7%) | Disease of Heart: 3 (10%) | Assault (Homicide): 1 (3%) | Diseases of Heart: 17 (4%) | Diseases of Heart: 36 (6%) | Chronic Liver Disease and Cirrhosis: 64 (7%) | Intentional Self-Harm (Suicide): 138 (8%) | Diabetes Mellitus: 224 (5%) | Chronic Lower Respiratory Diseases: 1974 (7%) | Cerebrovascular Diseases: 2349 (7%) |
| 6 | Septicemia: 2 (4%) | Cerebrovascular Diseases: 1 (3%) | Cerebrovascular Diseases: 1 (3%) | Congenital Malformations, Deformations, and Chromosomal Abnormalities: 8 (2%) | Chronic Liver Diseases and Cirrhosis: 21 (3%) | Diabetes Mellitus: 31 (4%) | Diabetes Mellitus: 72 (4%) | Chronic Liver Disease and Cirrhosis: 208 (5%) | Accidents (Unintentional Injuries): 1498 (6%) | Chronic Lower Respiratory Diseases: 2260 (6%) |
| 7 | Cerebrovascular Disease: 1 (2%) | Chronic Lower Respiratory Diseases: 1 (3%) | Chronic Lower Respiratory Diseases: 1 (3%) | Chronic Lower Respiratory Diseases: 7 (2%) | Diabetes Mellitus: 14 (2%) | Assault (Homicide): 24 (3%) | Cerebrovascular Diseases: 63 (4%) | Cardiovascular Diseases: 175 (4%) | Diabetes Mellitus: 1056 (4%) | Diabetes Mellitus: 1399 (4%) |
| 8 | Certain Conditions Originating in the Perinatal Period: 1 (2%) | *In Situ* Neoplasms, Benign Neoplasms, and Neoplasms of Uncertain or Unknown Behavior: 1 (3%) | Complications of Medical and Surgical Care: 1 (3%) | Influenza and Pneumonia: 4 (1%) | Cerebrovascular Diseases: 10 (2%) | Cerebrovascular Diseases: 21 (2%) | Chronic Lower Respiratory Diseases: 37 (2%) | Intentional Self-Harm (Suicide): 158 (4%) | Nephritis, Nephrotic Syndrome, and Nephrosis: 444 (2%) | Intentional Self-Harm (Suicide): 815 (2%) |
| 9 | Enterocolitis Due to *Clostridium Difficile*: 1 (2%) | Pneumonitis Due to Solids and Liquids: 1 (3%) | Disease of Heart: 1 (3%) | Legal Intervention: 3 (1%) | Legal Intervention: 6 (1%) | Chronic Lower Respiratory Diseases: 11 (1%) | Influenza and Pneumonia: 62 (2%) | Influenza and Pneumonia: 62 (2%) | Influenza and Pneumonia: 410 (2%) | Chronic Liver Disease and Cirrhosis: 682 (2%) |
| 10 | *In Situ* Neoplasms, Benign Neoplasms, and Neoplasms of Uncertain or Unknown Behavior: 1 (1.82%) | N/A | Influenza and Pneumonia: 1 (3%) | Diabetes Mellitus: 2 (1%) | Septicemia: 6 (1%) | Septicemia: 9 (1%) | Influenza and Pneumonia: 23 (1%) | Nephritis, Nephrotic Syndrome, and Nephrosis: 42 (1%) | Septicemia: 349 (1%) | Influenza and Pneumonia: 512 (2%) |

**Figure 1.**
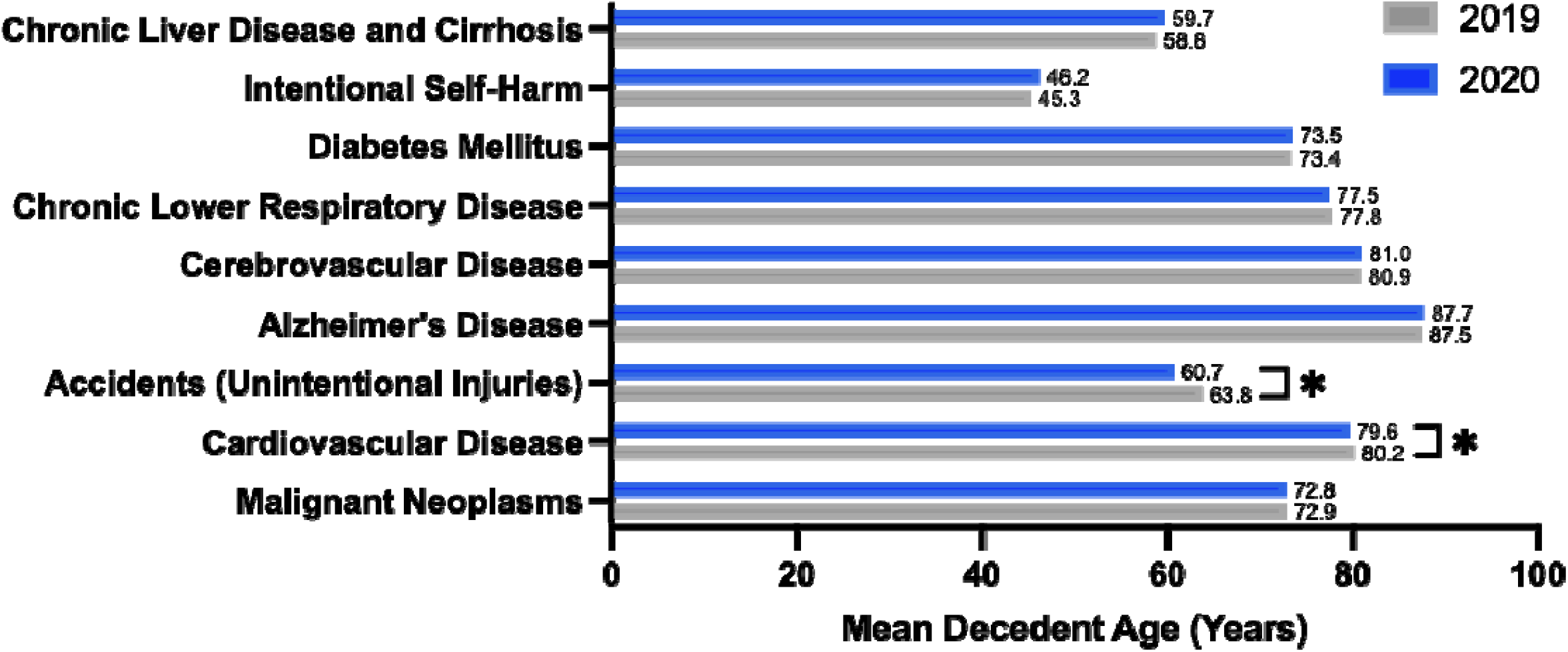
Mean Decedent Age for Leading Causes of Death in Minnesota, Stratified by Year.

### Cause of Death Dynamics Stratified by Gender

Minnesota reported 23,043 male resident decedents in 2019, and 26,629 in 2020, accounting for a significant 16% increase (95%, P < 0.001). Likewise, this increase in male resident decedents was paralleled by a significant 14% increase in female resident decedents, with 22,351 female resident decedents reported in 2019 and 25,563 reported in 2020 (95%, P < 0.001). However, the relative changes in male and female resident decedents between 2019 and 2020 were found to be statistically insignificant when compared to one another (95% CI, P = 0.4232) (Supplement Table 1).

The leading causes of death for male decedents in 2019 were malignant neoplasms, cardiovascular disease, and accidents or unintentional injuries (30%, 24%, and 9%, respectively). In 2020, the male mortality attributed to malignant neoplasms and cardiovascular disease decreased (25% and 22%, respectively) to accommodate for mortality attributed to COVID-19 (13%). When assessing the significance of these decreases, only changes in malignant neoplasms and cardiovascular disease were found to be significant (95% CI, P < 0.001). For females, the leading causes of mortality in 2019 were malignant neoplasms, cardiovascular disease, and Alzheimer’s disease (28%, 23%, and 10%, respectively). The mortality attributed to malignant neoplasms and cardiovascular disease decreased for females in 2020 (25% and 20%, respectively), as COVID-19 mortality was reported as 12% (Supplement Tables 2, 3). Only the changes in malignant neoplasms and cardiovascular disease were found to be significant (95% CI, P <0.001).

### Cause of Death Dynamics Stratified by Ethnicity

In 2019, Minnesota reported 41,585 non-Hispanic White resident decedents. In 2020, 47,121 non-Hispanic White decedents were reported, accounting for a significant 13% increase from 2019 (95% CI, P < 0.001). The leading causes of mortality for non-Hispanic White decedents in 2019 were malignant neoplasms, cardiovascular disease, and Alzheimer’s disease (29%, 24%, and 8%, respectively). In 2020, the mortality attributed to malignant neoplasms and cardiovascular disease decreased significantly to 25% and 22%, respectively (95% CI, P < 0.001).

1,592 non-Hispanic Black decedents were reported in 2019, and 2,129 were reported in 2020, constituting a significant 34% increase from 2019 (95% CI, P < 0.001). Non-Hispanic Black decedents possessed malignant neoplasms, cardiovascular disease, and accidents or unintentional injuries as their leading causes of mortality in 2019. The proportions of mortality attributed to malignant neoplasms and cardiovascular disease decreased significantly in 2020, to 20% and 15%, respectively (95% CI, P < 0.001).

Minnesota reported 516 Hispanic decedents in 2019, and 614 in 2020. The 19% increase in Hispanic decedents between 2019 and 2020 was significant (95% CI, P < 0.001). The top causes of death for Hispanic decedents in 2019 were reported as malignant neoplasms, accidents or unintentional injuries, and cardiovascular disease (28%, 17%, and 15%, respectively). All three forms of mortality decreased in prevalence in 2020, but only the decrease in malignant neoplasms was significant (95% CI, P = 0.019).

The COVID-19 decedents in Minnesota were disproportionately White, composing 86% of all COVID-19 decedents in 2020. However, disparities arose when examining the proportion of COVID-19 mortality within the total decedents based on race or ethnicity. Minnesota’s non-Hispanic White population possessed 4,393 COVID-19 decedents; and this composed 12% of their mortality for 2020. In contrast, Minnesota’s non-Hispanic Black and Hispanic populations possessed 262 and 117 COVID-19 decedents, which corresponded to 15% and 22% of their overall mortality, respectively (Supplement Tables 4, 5). Moreover, the leading cause of death for Minnesota’s Hispanic population in 2020 was COVID-19.

### Risk Factors for Acute Respiratory Distress Syndrome (ARDS) in Minnesota Decedents for 2020, Stratified by Ethnicity

Risk factors for ARDs, as described above, included select comorbidities and an age older than 65 years. When examining Minnesota’s non-Hispanic White COVID-19 decedents, 93% were older than 65 years, 8% had at least one comorbidity for ARDS listed as a contributing cause of death, and 7% had both an age and comorbidity risk factor. Within Minnesota’s non-Hispanic Black COVID-19 decedents, 68% were at or above 65 years old and 6% at least one comorbidity for ARDS listed as contributing cause of death. A similar trend was observed in Minnesota’s Hispanic COVID-19 decedent population, where 56% were at least 65 years old and 9% were reported to have at least one comorbidity for ARDS listed as a contributing cause of death. Minnesota’s Hispanic population had the greatest proportion of COVID-19 decedents under the age of 65, 44%, followed by the non-Hispanic Black and White COVID-19 decedents with 32% and 7%, respectively (Supplement Tables 6,7).

### Location as a Risk Factor for COVID-19 Mortality

Minnesota’s population is concentrated within the Twin Cities metropolitan area. In 2019, 22,790 decedents were reported in the Twin Cities. In 2020, the Twin Cities possessed 26,641 decedents, constituting a 13% significant increase from 2019 (95% CI, P < 0.001). Out of Minnesota’s 5,111 COVID-19 decedents in 2020, 3,055 or 60%, expired in the Twin Cities.

Another substantial location-based risk factor for COVID-19 mortality was nursing home residence due to increased potential for transmission and concentration of individuals with pre-existing comorbidities.^15-17^ In 2019, Minnesota reported 9,152 nursing home decedents and in 2020, there were 11,514 nursing home decedents, constituting a significant 26% increase in nursing home decedents over two years (95% CI, P < 0.001). In 2019, the primary causes of death for nursing home decedents were cardiovascular disease, Alzheimer’s disease, and malignant neoplasms (28%, 20%, and 15%, respectively). In 2020, COVID-19 became the leading cause of death, accounting for 23% of nursing home decedent mortality. Cardiovascular disease and Alzheimer’s disease shifted to second and third leading causes of death for nursing home decedents in 2020 at 20% and 16%, respectively (Figure 2). The average nursing home decedent age was 86.6 in 2019, and it decreased to 86.4 in 2020. However, the observed decrease in nursing home decedent age was found to be insignificant (95% CI, P = 0.057).

**Figure 2.**
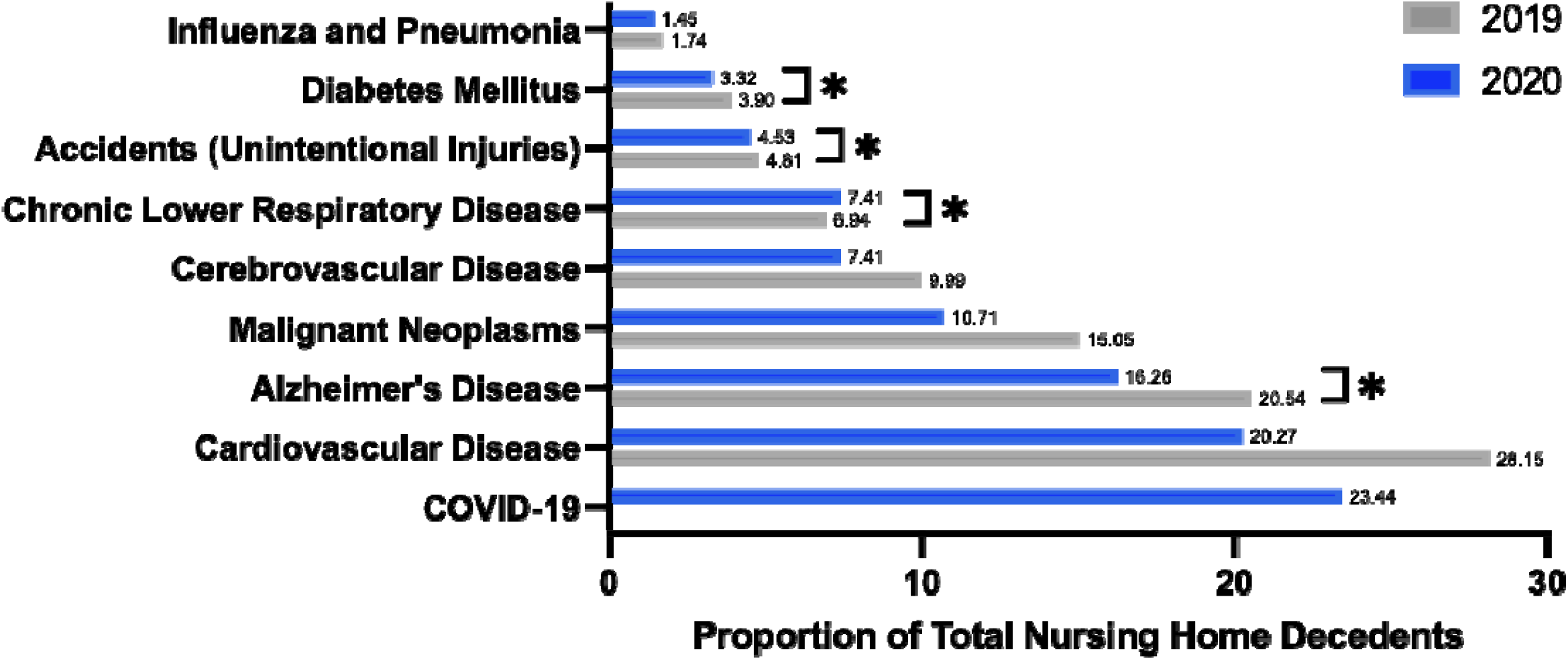
Top Causes of Death for Nursing Home Decedents in Minnesota, Stratified by Year.

### Location as a Risk Factor for COVID-19 Mortality, Stratified by Ethnicity and Gender

Most Minnesota nursing home residents are White and therefore, the non-Hispanic White population experienced a disproportionate risk for mortality during the COVID-19 pandemic.^18,19^ In 2020, 1,837 of COVID-19 decedents resided in a nursing home at the time of death. The quantity of non-Hispanic White nursing home decedents significantly increased from 8,870 individuals in 2019 to 11,147 individuals in 2020 (95% CI, P < 0.001). In contrast, only 130 non-Hispanic Black nursing home decedents were reported in 2019 and 136 in 2020. Hispanic decedents composed an even smaller share of nursing home decedents, with 45 reported in 2019 and 46 reported in 2020 (Supplement Table 8).

Furthermore, nursing home residents in Minnesota are overwhelmingly female due to the augmented life expectancy compared to males.^20,21^ As such, female nursing home residents were at an increased risk for COVID-19 mortality. In 2020, 1,086 COVID-19 nursing home decedents were female while only 751 were male. A significant difference in mortality was observed for female nursing home residents relative to male nursing home residents (95% CI, P < 0.001) (Supplement Table 9). Overall, female COVID-19 nursing home decedents composed 17% of all nursing home decedents and 42% of all White COVID-19 decedents in 2020.

Additional demographic mortality analysis pertaining to gender, age, ethnicity, and residence can be found in Supplement Tables 10-26.

## DISCUSSION

This retrospective study of mortality dynamics in Minnesota between 2019 and 2020 demonstrated that the emergence of the COVID-19 pandemic in 2020 (i) substantially altered mortality in Minnesota relative to the pre-pandemic period; and (ii) was associated with exacerbation of pre-existing racial and ethnic mortality disparities.

The augmented prevalence of racial and ethnic mortality disparities secondary to the COVID-19 pandemic has been well-reported in the literature, particularly with regard to Black and Hispanic populations.^22-26^ This study illustrates a parallel increase in racial and ethnic mortalities present in Minnesota during the onset of the COVID-19 pandemic in 2020, despite numerous public health initiatives aimed at limiting viral transmission and fatality in the Twin Cities Metro area where most individuals identifying as non-White reside.^27,28^ Additional research is warranted to identify factors that may have contributed to heightened racial and ethnic mortality disparities in 2020, such as workplace precautions and cooperation with public health measures.

Both Black and Hispanic individuals experienced disproportionate increases in mortality during 2020.This clearly reflected the pre-existing racial and ethnic health disparities in Minnesota as well as greater exposure to COVID-19 infection through both majority metropolitan residence and employment in essential industries, such as health care and social assistance, manufacturing, and accommodation and food services.^28,29^ Black and Hispanic COVID-19 decedents in Minnesota also expired at markedly younger ages relative to White COVID-19 decedents. The observed racial and ethnic mortality disparities also aligned with increased rates of hospitalization and death rates for Black and Hispanic individuals in 2020.^27-29^ These disparities reflect the poor social determinants of health in Minnesota, and the barriers to adequate healthcare and socioeconomic challenges, in part, put forth by generations of systemic racism.^30,31^

Of note, Minnesota nursing home residents also experienced substantial mortality disparities in 2020 secondary to housing conditions associated with higher rates of viral transmission and greater prevalence of comorbid conditions, including advanced age. Increased mortality disparities were observed for white female nursing home residents due to greater life expectancy.^20,21^

This study examined changes in mortality dynamics for Minnesota residents and excluded any individuals without state residency status at the time of death. Health disparities are fundamentally rooted in structural inequities including resources and infrastructure available in communities, such as affordable housing and economic development opportunities.^30^ Minnesota possesses some of the worst quality of life indices for non-White individuals in the entire United States, and respective health disparities during 2020 with the onset of the COVID-19 pandemic reflect the state’s racial and ethnic imbalance.^31^ Identifying geographic health disparities will assist in informing policy development in Minnesota and prioritize investment in initiatives combating health disparities in marginalized communities.

### Limitations

This study possesses several limitations. First, the study highlighted disparities reported for Black and Hispanic individuals during the COVID-19 pandemic due, in part, to occupation in essential industries but lacked reliable data regarding decedent occupation. Second, Hispanic decedents are historically underreported due to limited access to health services.^32,33^ It is thought that the reporting of Hispanic decedents in this study is incomplete to ambiguities in the collection of decedent race and ethnic information. Third, decedent data may be incomplete; data for all resident deaths in Minnesota were included, but a small quantity of unreported resident deaths, both in Minnesota and other states where residents expired, cannot be disregarded. Fourth, the results obtained by this study centered upon Minnesota resident decedents are not generalizable to other states. Lastly, this study did not examine any change in cause of death dynamics for American Indian and Alaskan Native populations, despite reports that both groups experienced racial mortality disparities during the COVID-19 pandemic.^34,35^

## CONCLUSIONS

This retrospective analysis of mortality data in Minnesota from 2019 to 2020 demonstrated an increase in mortality disparities during the emergence of the COVID-19 pandemic that disproportionately impacted Black and Hispanic residents.

## Data Availability

All data produced in the present study are available upon reasonable request to the authors.

## ABBREVIATIONS

COVID-19: coronavirus disease of 2019;
ARDS: acute respiratory distress syndrome

## ACKNOWLEDGEMENTS

This research was facilitated by the Minnesota Department of Health, which helped access the mortality data. The authors thank Cheri Denardo, MPH (Office of Vital Records, Minnesota Department of Health) and Neeti Sethi (Office of Vital Records, Minnesota Department of Health) for their important contributions. Neither of these individuals received compensation for their contributions.

This research was supported by the University of Minnesota grant #P01AG005842. The funders had no role in the design and conduct of the study; collection, management, analysis, and interpretation of the data; preparation, review, or approval of the manuscript; and decision to submit the manuscript for publication.

