## Supplemental Data for "Cause of Death by Race and Ethnicity in Minnesota Before and During the COVID-19 Pandemic, 2019-2020"

**Supplemental Material**

| Gender | 2019 | 2020 |
| --- | --- | --- |
| Male | 23,043 | 26,629 |
| Female | 22,351 | 25,563 |

Supplemental Table **1** – Analysis of Change in Decedents Between 2019 and 2020, Stratified by Gender

Female - Change Between 2019 and 2020 = p < 0.005

Male - Change Between 2019 and 2020 = p < 0.005

Difference Between Male and Female Decedent Change for 2019 and 2020 = p > 0.05

| Cause of Death | Prevalence |
| --- | --- |
| Malignant Neoplasms | 28.20% |
| Diseases of Heart | 23.20% |
| Alzheimer Disease | 10.40% |
| Cerebrovascular Diseases | 8.10% |
| Accidents (Unintentional Injuries) | 7.00% |
| Chronic Lower Respiratory Diseases | 6.70% |
| Diabetes Mellitus | 3.40% |
| Influenza and Pneumonia | 1.60% |
| Chronic Liver Disease and Cirrhosis | 1.60% |
| Nephritis, Nephrotic Syndrome, and Nephrosis | 1.50% |

Supplemental Table **2** – Top 10 Ranked Causes of Death for Females in Minnesota in 2019

p-value for accidents (unintentional injuries) < 0.005; everything else p > 0.05

| Cause of Death | Prevalence |
| --- | --- |
| Malignant Neoplasms | 24.50% |
| Diseases of Heart | 19.90% |
| COVID-19 | 12.20% |
| Alzheimer Disease | 9.10% |
| Cerebrovascular Diseases | 6.80% |
| Accidents (Unintentional Injuries) | 6.50% |
| Chronic Lower Respiratory Diseases | 5.80% |
| Diabetes Mellitus | 3.20% |
| Chronic Liver Disease and Cirrhosis | 1.80% |
| Influenza and Pneumonia | 1.60% |

Supplemental Table **3** – Top 10 Ranked Causes of Death for Females in Minnesota in 2020

p-value for accidents (unintentional injuries) < 0.005; everything else p > 0.05

| Total COVID-19 Hispanic Decedents | Total COVID-19 Black Decedents | Total COVID-19 White Decedents |
| --- | --- | --- |
| 117 | 262 | 4,393 |

Supplemental Table **4** – COVID-19 Decedents in Minnesota in 2020, Stratified by Ethnicity

| Ethnicity | COVID-19 Decedents | Relative Proportion of Total Decedents |
| --- | --- | --- |
| White | 4,393 | 11.93% |
| Black | 262 | 14.75% |
| Hispanic | 117 | 22.24% |

Supplemental Table **5**- Prevalence of COVID-19 As Leading Cause of Death in Minnesota in 2020, Stratified by Ethnicity

| Risk Factors  for ARDS | Hispanic COVID-19 Decedents | Black COVID-19 Decedents | White COVID-19 Decedents |
| --- | --- | --- | --- |
| Age > 65 | 65 (56.0%) | 179 (68.3%) | 4096 (93.2%) |
| Comorbidity Present* | 10 (8.5%) | 15 (5.7%) | 351 (8.0%) |
| Age > 65 & Comorbidity Present | 7 (6.0%) | 4 (1.5%) | 320 (7.3%) |

Supplemental Table **6** – Analysis of Risk Factors for Acute Respiratory Distress Syndrome in Minnesota Decedents for 2020, Stratified by Ethnicity

Comorbidities for ARDS Were Broadly Defined as Diabetes Mellitus, Hypertension, COPD, Asthma, Sepsis, and Malignancy

| Ethnicity | COVID-19 Decedents with Age < 65 |
| --- | --- |
| White | 297 (6.8%) |
| Black | 83 (31.7%) |
| Hispanic | 51 (44.0%) |

Supplemental Table **7** – COVID-19 Decedents in Minnesota in 2020 with Age > 65, Stratified by Ethnicity

| Ethnicity | Total Nursing Home Decedents in 2019 | Total Nursing Home Decedents in 2020 |
| --- | --- | --- |
| Hispanic | 45 | 46 |
| Black | 130 | 136 |
| White | 8,870 | 11,147 |

Supplemental Table **8** – Change in Nursing Home Decedents in Minnesota Between 2019 and 2020, Stratified by Ethnicity

White: p < 0.005, Black: p = 0.60, Hispanic: p = 0.88

| Total White Nursing Home COVID-19 Decedents | White Female Nursing Home COVID-19 Decedents | White Male Nursing Home COVID-19 Decedents |
| --- | --- | --- |
| 1,837 | 1,086 (59.1%) | 751 (39.9%) |

Supplemental Table **9** – White COVID-19 Nursing Home Residents in 2020, Stratified by Gender

p-value < 0.001

(Females are 16.5% of All NH Decedents, 41.8% of All White COVID-19 Decedents)

| Ethnicity | COVID-19 Decedents in Twin Cities Metro Area | Total Decedents in Twin Cities Metro Area for 2020 |
| --- | --- | --- |
| Hispanic | 96 (22.1%) | 434 |
| Black | 248 (13.0%) | 1,913 |
| White | 2,428 (10.7%) | 22,682 |

Supplemental Table **10** – Total Decedents and COVID-19 Decedents in Minnesota in 2020, Stratified by Ethnicity and Residence in Twin Cities Metro Area

| Cause of Death | White P-Value | Black P-Value | Hispanic P-Value |
| --- | --- | --- | --- |
| Malignant Neoplasms | 1.963908e-35 | 0.009396394 | 0.01875292 |
| Diseases of the Heart | 3.288256e-18 | 0.0003628725 | 0.05215226 |
| Accidents (Unintentional Injuries) | 0.7936672 | 0.1250501 | 0.8711349 |
| Alzheimer’s Disease | 6.108231e-07 | 0.7126529 | 0.5600408 |
| Cerebrovascular Disease | 3.960718e-11 | 0.9568376 | 0.3657008 |
| Chronic Lower Respiratory Disease | 6.009057e-09 | 0.4076231 | 0.1667076 |
| Diabetes Mellitus | 0.002080636 | 0.6464613 | 0.4660813 |
| Self-Harm (Suicide) | 3.723821e-05 | 0.02333679 | 0.2578004 |
| Chronic Liver Disease and Cirrhosis | 0.03810427 | 0.3875851 | 0.4246029 |

Supplemental Table **11** - Top Ranked Overlapping Causes of Death and Respective Significance Values Between 2019 and 2020, Stratified by Ethnicity

| *Cause of Death* | *Male P-Value* | *Female P-Value* |
| --- | --- | --- |
| Malignant Neoplasms | *1.182389e-22* | *1.364912e-14* |
| Diseases of the Heart | *5.552567e-08* | *5.759803e-12* |
| Accidents (Unintentional Injuries) | *0.1678201* | *0.04511007* |
| Alzheimer’s Disease | *0.001250274* | *6.459593e-05* |
| Cerebrovascular Disease | *4.573866e-06* | *1.363318e-06* |
| Chronic Lower Respiratory Disease | *9.249092e-08* | *0.003721146* |
| Diabetes Mellitus | *0.007512666* | *0.122243* |
| Self-Harm (Suicide) | *0.001246719* | *0.01141841* |
| Chronic Liver Disease and Cirrhosis | *0.4737003* | *0.01484652* |

Supplemental Table **12** - Significance of Gender for Change in Overlapped Ranked Cause of Death in White Population Between 2019 and 2020

| *Cause of Death* | *Male P-Value* | *Female P-Value* |
| --- | --- | --- |
| Malignant Neoplasms | *0.002830498* | *0.6900492* |
| Diseases of the Heart | *0.006016032* | *0.02894163* |
| Accidents (Unintentional Injuries) | *0.04967029* | *0.812716* |
| Alzheimer’s Disease | *0.875752* | *0.7689449* |
| Cerebrovascular Disease | *0.6473081* | *0.6234047* |
| Chronic Lower Respiratory Disease | *0.3393505* | *0.9977864* |
| Diabetes Mellitus | *0.1182777* | *0.297905* |
| Self-Harm (Suicide) | *0.09274243* | *0.1262396* |
| Chronic Liver Disease and Cirrhosis | *0.7204145* | *0.9977864* |

Supplemental Table **13** – Significance of Gender for Change in Overlapped Ranked Cause of Death in Black Population Between 2019 and 2020

| *Cause of Death* | *Male P-Value* | *Female P-Value* |
| --- | --- | --- |
| Malignant Neoplasms | *0.1241784* | *0.1280676* |
| Diseases of the Heart | *0.08779004* | *0.4227882* |
| Accidents (Unintentional Injuries) | *0.7452309* | *1.0* |
| Alzheimer’s Disease | *0.09905213* | *0.7995423* |
| Cerebrovascular Disease | *0.5516514* | *0.736018* |
| Chronic Lower Respiratory Disease | *0.4135052* | *0.4990365* |
| Diabetes Mellitus | *0.1326572* | *0.7026158* |
| Self-Harm (Suicide) | *0.4039221* | *0.5467953* |
| Chronic Liver Disease and Cirrhosis | *0.6204759* | *0.8394417* |

Supplemental Table **14** – Significance of Gender for Change in Overlapped Ranked Cause of Death in Hispanic Population Between 2019 and 2020

| Mean Decedent Age – 2019 | Mean Decedent Age – 2020 |
| --- | --- |
| 75.8 | 75.8 |

Supplemental Table **15** – Change in Mean Decedent Age Between 2019 and 2020

p-value > 0.05

| Cause of Death | Mean Decedent Age – 2019 | Mean Decedent Age – 2020 |
| --- | --- | --- |
| Malignant Neoplasms | 72.9 | 72.8 |
| Cardiovascular Disease | 80.2 | 79.6 |
| Accidents (Unintentional Injuries) | 63.9 | 60.7 |
| Alzheimer’s Disease | 87.5 | 87.7 |
| Cerebrovascular Disease | 80.9 | 81.0 |
| Chronic Lower Respiratory Disease | 77.8 | 77.5 |
| Diabetes Mellitus | 73.4 | 73.5 |
| Intentional Self-Harm (Suicide) | 45.3 | 46.4 |
| Chronic Liver Disease and Cirrhosis | 59.7 | 58.8 |

Supplemental Table **16** – Analysis of Change in Decedent Age for Top Overlapping Ranked Causes of Death, Stratified by Year

Cardiovascular Disease = p < 0.005

Accidents (Unintentional Injuries) = p < 0.005

| Decedents in Minnesota for 2019, Excluding COVID-19 Decedents | Decedents in Minnesota for 2019, Excluding COVID-19 Decedents |
| --- | --- |
| 45,396 | 47,081 |

Supplemental Table **17** – Change in Decedents in Minnesota Between 2019 and 2020, Excluding COVID-19 Mortality in 2020

p-value < 0.005

| Total Decedents in Minnesota for 2019 | Total Decedents in Minnesota for 2020 |
| --- | --- |
| 45,396 | 52,192 |

Supplemental Table **18** – Change in Decedents in Minnesota Between 2019 and 2020, Including COVID-19 Mortality in 2020

p-value < 0.05

| Year | Total Hispanic Decedents in Minnesota | Total Black Decedents in Minnesota | Total White Decedents in Minnesota |
| --- | --- | --- | --- |
| 2019 | 516 | 1,592 | 41,585 |
| 2020 | 614 | 2,129 | 47,121 |
| Significance Testing | p-value = 3.653e-05 | p-value < 2.2e-16 | p-value < 2.2e-16 |

Supplemental Table **19** – Change in Decedents in Minnesota, Stratified by Year and Ethnicity

| Year | Decedents in Twin Cities Metro Area | Non-COVID-19 Decedents in Twin Cities Metro Area |
| --- | --- | --- |
| 2019 | 22,790 | 22,790 |
| 2020 | 26,641 | 23,596 |

Supplemental Table **20** –Total Decedents in Minnesota in Twin Cities Metro Area, Stratified by Year and Inclusion of COVID-19 Decedents for 2020

Change between 2019 and 2020 for decedents in Twin Cities Metro: p < 0.005

Change between 2019 and 2020 for non-COVID-19 decedents in Twin Cities Metro: p < 0.005

| COVID-19 Decedents for 2020 | COVID-19 Decedents in the Twin Cities Metro Area for 2020 |
| --- | --- |
| 5,111 | 3,055 (59.8%) |

Supplemental Table **21** – COVID-19 Decedents in Minnesota in 2020, Stratified by Residence in Twin Cities Metro Area

| Total Nursing Home Decedents in 2019 | Total Nursing Home Decedents in 2020 |
| --- | --- |
| 9,152 | 11,514 |

Supplemental Table **22** – Change in Nursing Home Decedents in Minnesota Between 2019 and 2020

p-value < 0.005

| Average Nursing Home Decedent Age in 2019 | Average Nursing Home Decedent Age in 2020 |
| --- | --- |
| 86.6 | 86.4 |

Supplemental Table **23** – Average Nursing Home Decedent Age in Minnesota, Stratified by Year

p = 0.23

**P-Value for Nursing Home Decedents and Ethnicity**

*Significant Difference in Decedents of One Ethnicity vs. Others:*

White Relative to Non-White: p < 0.05

Black Relative to Non-Black: p < 0.05

Hispanic Relative to Non-Hispanic: p < 0.05

| Ethnicity | Decedents in 2019 | Decedents in 2020 | Relative Percent Change |
| --- | --- | --- | --- |
| White | 41,585 | 47,121 | 13% |
| Black | 1,592 | 2,129 | 34% |
| Hispanic | 516 | 614 | 19% |

Supplemental Table **24** – Relative Percent Change in Decedents in Minnesota, Stratified by Year and Ethnicity

p-values < 0.005

| Cause of Death | Proportion of Total Male Decedents |
| --- | --- |
| Malignant Neoplasms | 29.50% |
| Diseases of Heart | 24.40% |
| Accidents (Unintentional Injuries) | 9.00% |
| Chronic Lower Respiratory Diseases | 6.20% |
| Cerebrovascular Diseases | 5.40% |
| Diabetes Mellitus | 4.50% |
| Alzheimer Disease | 4.40% |
| Intentional Self-Harm (Suicide) | 3.40% |
| Chronic Liver Disease and Cirrhosis | 2.30% |
| Nephritis, Nephrotic Syndrome, and Nephrosis | 1.50% |

Supplemental Table **25** – Top 10 Ranked Causes of Death for Males in Minnesota in 2019

p-value for accidents (unintentional injuries) < 0.005; everything else p > 0.05

| Cause of Death | Proportion of Total Male Decedents |
| --- | --- |
| Malignant Neoplasms | 24.50% |
| Diseases of Heart | 21.50% |
| COVID-19 | 12.60% |
| Accidents (Unintentional Injuries) | 9.60% |
| Chronic Lower Respiratory Diseases | 4.90% |
| Cerebrovascular Diseases | 4.40% |
| Diabetes Mellitus | 3.90% |
| Alzheimer Disease | 3.70% |
| Intentional Self-Harm (Suicide) | 2.70% |
| Chronic Liver Disease and Cirrhosis | 2.70% |

Supplemental Table **26** – Top 10 Ranked Causes of Death for Males in Minnesota in 2020

*p-value for accidents (unintentional injuries) < 0.005; everything else p > 0.05*
